# Perceptions of children and young people in England on the smokefree generation policy: a focus group study

**DOI:** 10.1101/2024.04.29.24306422

**Authors:** Nathan P Davies, Rachael L Murray, Tessa Langley, Joanne R Morling, Manpreet Bains

## Abstract

**Background:** This study investigates perceptions of young people towards the planned smokefree generation (SFG) policy in England, which will ban sale of tobacco products to those born in or after 2009. It focuses on SFG policy acceptability, design and implementation.

**Methods:** We conducted 7 semi-structured focus groups with 36 participants aged 12 - 21 (mean = 15) in England over video call and in person. 21 participants were female and 15 male. Participants were purposively sampled to include those from areas of greater deprivation and for use of tobacco or e-cigarettes. Data was analysed using the framework approach.

**Results:** Participants expressed broadly negative perceptions towards tobacco and its manufacturers. Most participants supported SFG policy goals and its focus on freedom from addiction and harm. Many believed the law would benefit from stringent enforcement, inclusion of e-cigarette products, tobacco licensing, and input from young people. A minority raised concerns about the loss of freedom to purchase tobacco and believed it would have little effect on smoking rates.

**Conclusion:** Communication of the freedom-giving nature of SFG is likely to resonate with many young people. Enforcement, communication, and involvement of young people in SFG should be considered carefully to maximise policy impact.

**What is already known on this topic:** Observational and modelling studies of raising the legal age of sale of tobacco show its effectiveness in reducing smoking rates in target populations. The UK is currently on track to be the first nation to introduce a generational ban on tobacco products, but little is known about young people’s perceptions on this policy.

**What this study adds:** The overarching goal and preventative approach of SFG has the power to resonate with young people, including nicotine product users, but there is likely to be a small minority opposed to SFG on philosophical principles and perceptions of limited effectiveness. Young people may have mixed feelings about e-cigarettes being excluded from SFG due to misperceptions of equivalent harm between products.

**How this study might affect research, practice or policy:** Our study suggests involvement of young people in SFG’s design and accompanying communication is likely to strengthen its legitimacy and appeal. Our sample were largely supportive of well-resourced, consistent enforcement of SFG law with strong penalties for retailers who break the law.

## INTRODUCTION

Preventing smoking initiation and addiction in children and young adults has long been crucial to the reduction of smoking rates.[1] Evidence from the first COVID-19 lockdown in England found a 25% increase in smoking prevalence among 18–34-year-olds.[2] This indicates that long-term decline in smoking in young adults in countries with strong tobacco control should not be taken for granted.

One policy option for reducing smoking amongst children and young adults is a minimum legal age of sale for tobacco products. The WHO Framework Convention on Tobacco Control (FCTC) indicates members who have signed should restrict tobacco sales to 18, or the age of adulthood set under local law.[3] The FCTC exhorts signatories to go beyond its minimum requirements, paving the way for minimum legal ages of sale above 18. The United States has implemented a federal law setting the minimum legal sale age at 21 (Tobacco 21) [4] and a small number of other nations have been reported to have introduced a minimum legal sale age of at least 20.[5] A systematic review of such laws included nineteen studies, solely from the United States. It found the introduction of Tobacco 21 was likely to reduce smoking rates, particularly for older school-aged groups and those aged 18-20.[6] However, there were differential effects by place of implementation. For example, impact of the policy was less clear for the state of California[7–9] which had issues with enforcement of the law immediately after implementation and several special exemptions.[10] Qualitative research into Tobacco 21 suggests that while some young people are supportive of the policy, some disagree with the way it appears to subvert the traditional age of adulthood and doubt its effectiveness.[11,12]

There are other age-related policy options. A smoke-free generation policy (SFG), also known as a tobacco-free generation policy, bans the sale of tobacco products to anyone born after a specific year. Its advocates argue that it overcomes a key limitation of age-based age-of-sale policies, which is that they convey a message that tobacco use is socially acceptable and perhaps even a rite of passage into adulthood.[13–15] Simulation modelling for New Zealand[16,17] Singapore[18,19] and the United Kingdom[20] all find that, over long timeframes, an SFG policy is likely to be one of the most effective tobacco control policies for reducing smoking rates in children and young people.

Several areas have proposed variations on SFG policies, including Tasmania (Australia), Finland, Malaysia and Denmark.[21–24] However, these proposals have foundered under changes of Government, political changes of heart, or the possibility or reality of legal challenges. The small town of Brookline (USA) and the city of Balanga (Philippines) continue to fight legal battles to maintain implementation of their SFG policies.[25,26]

New Zealand’s SFG policy, which was to be introduced alongside denicotinisation and a reduction in outlets selling tobacco, was halted in late 2023. The newly elected government argued its repeal would lead to increased tax intake to fund separate tax cuts.[27]

On 4 October 2023, the UK Government announced it would be implementing an SFG model that banned the sale of tobacco to those born on or after 1 January 2009.[28] The Government’s intention is for the law to cover a variety of tobacco products, but not electronic cigarettes (e-cigarettes), for which different restrictions are being introduced, including a ban on disposable e-cigarettes.[29] Sixty-seven per cent of adults in the UK general public[30] and 63.2% of respondents to the Government’s public consultation supported the idea.[31] Support amongst under-18s has not been studied.

To assess acceptability and inform SFG implementation, it is important to study the perceptions of young people, particularly those who will be directly impacted by the policy. We conducted focus groups with a range of young people who either will be directly affected by SFG in England, or who were at an age where they could conceptualise SFG affecting themselves and their peers. We sought to understand their views on the SFG proposals in England.

## METHODS

### Design and epistemological approach

Given the inherently social nature of youth smoking, the research was underpinned by a constructionist approach. Participants’ words were given meaning and interpreted through the context of their social and wider environments.[32] The lead researcher, ND, a male public health specialist with experience in qualitative research, undertook bracketing [33] to identify and mitigate prior assumptions on both population and policy. The COREQ framework has been used to support transparent reporting (Supplementary file 1).[34]

### Sampling and recruitment

Our inclusion criteria incorporated children and young people living in England aged between 11-21, who were those who would be directly affected by SFG, and older children and young people who would not be affected but who could conceive of the law affecting their age group. We began by sampling for maximum variation across age, gender, ethnicity and region, before purposively sampling for theoretical saturation of groups that were underrepresented in our data, such as certain age groups or tobacco or e-cigarette use. We purposively sampled for the majority of participants to live in areas of greater deprivation to enable consideration of those most affected by health inequalities.

We recruited participants through the support of a range of organisations working with children and young people, such as schools, colleges, youth groups, and education and employment charities. These organisations purposively approached young people based on our inclusion criteria. Participant information sheets were provided in advance by gatekeeper organisations and the lead interviewer also verbally discussed them with participants ahead of focus groups. Participants completed an online or paper consent form before focus groups commenced.

### Data collection

Participants completed a paper or online questionnaire reporting demographic details any smoking and e-cigarette history. Based on prior literature[35], the UK government’s SFG command paper[36] and the COM-B framework[37] we developed a topic guide (Supplementary file 2) covering (1) personal experiences of tobacco and e-cigarette use (2) the concept of SFG (3) the implementation of SFG. Participants were offered a £10 shopping voucher to compensate for their time.

ND conducted focus groups in-person and over MS Teams, depending on organisational and participant preference. For safeguarding reasons, an adult from gatekeeping organisations was present for focus groups with participants under the age of 18. They sat away from the main group or had their camera switched off, and were asked not to speak. Focus groups lasted between 26 – 36 minutes (mean = 32 minutes) and were recorded via MS Teams or a handheld recording device. Initial transcription was made through the University of Nottingham’s automated transcription service and corrections made by ND. No significant new themes or ideas were noted in the sixth or seventh focus groups and so no further recruitment took place.

### Data analysis

Transcripts were read several times to support data familiarisation. The framework approach was used to guide analysis.[38] Initial codes, themes, and subthemes were generated inductively by ND with the use of NVivo 12.[39] A sample of transcripts were double-coded by MB to provide triangulation and enhance the credibility of the analysis. [40]. An initial analytical framework was developed to index and chart data with the incorporation of a constant comparative method to ensure all data was considered.[41]. The framework was reviewed between ND and MB at various points in the analysis before being finalised.

### Ethical approval

Ethical approval was granted by the Faculty of Medicine and Health Sciences Research Committee at the University of Nottingham (reference FHMS 39-1023).

### Public involvement

Three separate groups of public advisors aged 12-21 (n = 18) provided advice on inclusion criteria, recruitment methods and the topic guide.

## RESULTS

Seven focus groups were conducted comprising 36 participants aged 12-21 from across England. Some participants made an informed decision not to share their postcode or other personal details. Demographic details are reported in Table X. 58% of the sample were female, 47% were aged 12 – 14 and 61% were White British. 53% of the sample had tried tobacco and 82% had tried e-cigarettes. 59% of participants were from the most deprived quintile of English postcodes.

**Table 1:** Participant demographics.

| **Characteristics** | **n** |
| --- | --- |
| **Gender** |  |
| Female | 21 |
| Male | 15 |
| **Age (years)** |  |
| 12 – 14 | 17 |
| 15 – 17 | 14 |
| 18 – 21 | 5 |
| **E-cigarette use** |  |
| Never | 6 |
| Tried once or twice | 3 |
| Current use | 22 |
| Missing | 2 |
| **Tobacco use** |  |
| Never | 16 |
| Former use | 9 |
| Tried once or twice | 9 |
| Current use | 7 |
| Missing | 2 |
| **Postcode deprivation quintile (1 = most deprived)** |  |
| 1 | 10 |
| 2 | 7 |
| 3 | 7 |
| 4 | 2 |
| 5 | 3 |
| Missing | 7 |
| **Ethnicity** |  |
| White British | 22 |
| Gypsy Roma | 4 |
| Mixed background | 4 |
| Black | 2 |
| White Other | 1 |
| Any other background | 1 |
| Missing | 3 |

### Thematic framework

Four themes relating to the SFG policy were generated. The themes are: (1) Broad perceptions of tobacco and other nicotine products (2) Principles underpinning SFG (3) Impact of SFG and (4) Implementation of SFG. We identified some differences according to use of tobacco or e-cigarette products and some difference between age groups, but we did not identify differences between genders. Quotes signify gender, age, and smoking and e-cigarette status (T = tried, F = former, R = regular, N = never, S = smoked tobacco, V = vapes/e-cigarettes).

### Broad perceptions of tobacco and other nicotine products

Most participants, including the majority of current users of e-cigarettes and/or tobacco, had a negative view of tobacco and the tobacco industry, especially towards the industry’s methods of attracting young purchasers. Both those who had tried cigarettes and those who hadn’t expressed physical revulsion towards smoked tobacco, with just one participant expressing positive sentiment.

> *Eughh, fags are vile. (M17, TS/RV, FG2)*

Participants also expressed significant concerns around the health risks of e-cigarettes, largely centred around lack of knowledge of future damage. Participants were largely unconvinced that e-cigarettes were safer than tobacco, despite largely being aware that it was promoted as a quit aid for adults.

> *Thing is with vaping, it’s still quite new. So you don’t know the long-term effects of it. (F20, RV/TS, FG7)*

Younger participants had incomplete knowledge of existing age-of-sale laws, with many suggesting that the age of sale for cigarettes was 21 or that the age of sale for e-cigarettes was 16. For those who had tried or used tobacco or e-cigarettes whilst under the age of sale, there was a broad perception that they were relatively straightforward to access whilst underage. Methods ranged from supply or theft from older friends and family members, picking products up from the street and direct purchase from retailers.

> *When I was ten, my sisters’ ex-boyfriend. Yeah, he got me. He approached me to smoke cigarettes and start vaping and then I got addicted from there. (M14, TS/RV, FG3)*

### Principles underpinning SFG

Most participants supported the goals of SFG in its broadest sense. They conveyed a sense of enthusiasm towards the idea of being part of a smokefree generation.

> *I think it’s a nice thing to think about when people look back into the past they’ll think, wow, these people stopped smoking. (F13, NS/NV, FG1)*

Linked to this view was personal experience of the harm of tobacco. Several participants expressed feeling helpless about the harms that smoking exacted on their families, and several expressed how difficult it was to quit e-cigarettes or smoking and that the feeling of addiction took a toll on their lives.

> *But then when you think about it, it’s like don’t bother with (vaping) because then it’s gonna hurt me even more, don’t die from it. Like my grandmother died from cancer about 5 months ago, I keep on worrying about that, but then like I keep on forgetting about it. But like I go on and off (vapes), but then it hurts, to think. (M14, NS/RV, FG3)*

Several participants expressed the idea that SFG supported positive freedom *from* addiction to tobacco, and in doing so protected them or others from making a decision they would later struggle to reverse.

> *It’s a good idea. Because young people can get addicted easily. At the age of 18, it’s too hard for them to make a decision. So I think it’s good for them not to smoke at all. (M18, NS/TV, FG2)*

A small minority of participants, mostly those who currently smoked tobacco or used e-cigarettes, suggested SFG would jeopardise personal liberties, bodily autonomy or contravene the convention of 18 being the age of adulthood. Another small minority of participants, who were current users of e-cigarettes, expressed views of “not caring” about SFG because they had little interest in smoking tobacco and would only consider using e-cigarettes. This appeared to be linked to the conception of tobacco being “disgusting”, the relative cheapness of e-cigarettes and the attractiveness of e-cigarette flavouring, packaging and taste, although perceived safety was not raised.

Other participants often expressed confusion that e-cigarettes were not being included in the smokefree generation proposal, given what they perceived to be the greater problem of e-cigarette use amongst young people and potential equivalent health harms. Two participants felt SFG may lead to greater e-cigarette uptake.

> *(Vapes) have still got a really addictive component that you can have, and then you have a cigarette and it could feel the same. So it could still lead you onto it. So I think if you’re going to be banning tobacco products, you should ban nicotine products as well. (F21, RS/RV, FG7)*

### Impact of SFG

Participants were nuanced in their discussions of how effective SFG would be in reducing smoking rates, rarely dismissing it completely or predicting total success. The minority of participants against SFG on philosophical grounds were typically very sceptical of the effectiveness of SFG. Some expressed reservations the policy could backfire, suggesting that making tobacco sales illegal would increase its appeal amongst young people and lead to increased smoking rates and law-breaking.

> *It’s just gonna cause more illegal activity because there’s more people giving them illegally. So there’s no point. I don’t think there’s any point of doing anything with the cigarettes or the vapes because it’s just going to cause more havoc for the government. (F14,TS/RV, FG1)*

Several drew upon personal experiences of underage purchasing to envisage that retailers and young people (including themselves) would find a way round the regulations through parents or by identifying retailers happy to break the rules.

> *Once you see the packet of cigarettes on the table, and then the parents go somewhere, they will take it just like I do. (F14, RS/RV, FG6)*

However, many of the focus group conversations converged on a broad agreement that SFG would prevent young people from taking up tobacco smoking, but not prevent those who were already addicted.

### Implementation of SG

Many participants argued that retailers who breached the rules should face the harshest possible penalties, including jail sentences. There was considerable worry that some retailers would seek to bypass the law by existing methods such as covert under-the-counter sales.

There was also strong support for the idea of licensing tobacco retailers and reducing the number of outlets who could sell cigarettes.

> *In [area] you can buy anything… Just go to shop, say I want that, they give it to you. They don’t ask for nothing. I recommend you just shut down every shop in [area]. M15, FS/RV, FG6*

There was a plurality of perspectives on how the SFG should be supported by government communications. This ranged from a strong focus on the smokefree vision, to a strong focus on the health impact of tobacco, to the idea that significant government communication could backfire because young people would rebel against the perception of being told what to do.

There was strong agreement that children and young people should be involved in decision-making and implementation of SFG, especially in the design of the communications and providing insight into how tobacco is currently obtained underage. This was justified as being both the right thing to do and to provide unique perspectives into the lives of their peers.

> *I think (youth involvement) is a really good and important idea because I think it’s important that young people can sort of feel they are the smoke free generation. (M17, NS/NV, FG7)*

## DISCUSSION

### Summary of findings

Our findings offer a rich insight into the perspectives of children and young people who would be directly affected by the SFG law, or offered a young person’s perspective on the law, including strong representation of participants living in areas of higher deprivation and those with experience of tobacco use or e-cigarette use.

We found that many participants of all demographics and experiences with tobacco and e-cigarettes expressed revulsion towards tobacco products and manufacturers. These views were linked with a positive conception of the vision and aims of the smokefree generation and positive freedom from addiction, similar to findings from 17- and 18-year-olds in New Zealand on SFG.[35] The broad support for reducing youth smoking is reflective of findings in qualitative studies of male perceptions of Tobacco 21 in Singapore,[11] perceptions of vulnerable 13–19-year-olds of Tobacco 21 in Tasmania,[42] and perceptions of SFG in New Zealand.[35]

Many participants who were in general support of SFG still felt that, as a standalone policy, it would be insufficient to completely eradicate smoking in young people and strongly advocated for licensing for a smaller number of tobacco retailers, greater enforcement, and stronger penalties for errant retailers. Young people’s instincts and lived experience align with the evidence. The effect of age-of-sale bans for tobacco and other restrictions on availability of product is strongly linked to the degree of enforcement and awareness of the bans.[43,44] Recent modelling for New Zealand projected that licensing restrictions would have an early step-change in smoking.[17] Support for licensing restrictions is strongly articulated by young people, who are most affected by retailers that choose to sell products to those under the legal age of sale.

A small minority, largely those with a history of tobacco use, were strongly against the principle of restricting choice through SFG because of its infringement on individual freedom, and to a lesser extent, the subversion of the convention of adulthood beginning at 18. Similar findings were reported by some participants in qualitative studies in the United States, Singapore, Tasmania, and New Zealand.[11,35,42,45] Some participants also raised the issue of generational smoking and proxy purchasing. However, it appeared that participants were mainly considering the early years of the policy, where a 20-year-old born in 2007 could buy products for 18-year-olds born in 2009. As time progresses, the age difference between those who can be legally sold tobacco and those who cannot will steadily increase under SFG, a point made by several other participants.

Whether e-cigarettes should be covered by SFG mandates is a topic of debate. Our sample were largely supportive of e-cigarettes being covered by SFG and many found their omission the most illogical element of the UK government’s approach to SFG. This contrasts with findings from a qualitative study of 15 – 21 year olds with experience of e-cigarettes in US, where participants expressed disappointment that Tobacco 21 would cover e-cigarette products.[45] Participant views appeared to be partly driven by the idea that e-cigarettes are at least as harmful as cigarettes – a view known to be growing in the UK [46] - and partly by awareness of the far greater prevalence of e-cigarette use than tobacco use amongst young people in the UK. The UK Government have elected to ban disposable e-cigarettes and restrict its marketing and availability rather than include it as part of the SFG ban on tobacco sales in order to retain e-cigarettes as a quit aid for adults whilst reducing its appeal for children.[31] This is a relatively complex message to get across to the general public and our study shows that this is unlikely to be readily understood by young people. The incomparable harm caused by tobacco may need to be re-emphasised in Government communications.

### Policy implications

Our findings highlight two conditional patterns of support for SFG as a policy.[37] Firstly, most participants expressed excitement or support for the vision of a smokefree generation, but were keen that it was sufficiently enforced. Secondly, participants were very supportive of including young people in the decision-making process. This may guide approaches to both SFG policy implementation and government communication of SFG. Tobacco control policies should always seek to reduce health inequalities wherever possible. Participants living in areas of higher deprivation were all very aware of shops in their area who sold underage tobacco and e-cigarette products, referring to this phenomenon with an apparent sense of resignation. Raising the age of sale of tobacco to both 18[47] and 21[6] have been associated with reducing socioeconomic health inequalities. Governments introducing SFG policies should direct resources for enforcement and communication into areas with high youth smoking rates and lower adherence with existing age-of-sale laws in order to have the greatest impact on health inequalities.

Participants were generally enthusiastic about the idea of a license for tobacco that limited the number of retailers, one of the three pillars of the New Zealand smokefree legislation to be halted, [27] and a policy under consideration in the United Kingdom. This is a potential area for future exploration.

### Limitations

Our study has some weaknesses. We cannot extrapolate findings to the general population from our sample due to its qualitative nature. However, the depth of findings from a variety of areas suggests even many of those at greatest risk of tobacco smoking are supportive of the principles of the smokefree generation approach and take a positive view of freedom as protection from addiction and harm. The research also took place at a time where the UK policy landscape on e-cigarettes and tobacco was changing quickly. In the middle of data collection, the government announced plans to restrict e-cigarettes, including for disposable e-cigarettes to be banned. Researchers sought to ask questions in a similar manner and to avoid explaining any new laws to maintain consistency across focus groups and participant perspectives; however, knowing that this law was to be introduced may have influenced some of the discussion around inclusion or exclusion of e-cigarettes from SFG.

### Conclusions

These findings provide crucial evidence as to how young people, particularly those likely to be affected by the SFG policy, view its legitimacy, its likelihood of success and its optimal implementation. Participants were largely supportive of SFG, with a minority opposed, although many felt significant efforts would be required to enforce it. Our study suggests that the support of young people can be strengthened by including them in its design and implementation, re-emphasising the unique harms of tobacco, focusing on the big-picture SFG goal of a generation who does not smoke tobacco, and by putting time and resources into enforcement of retailers.

## Data availability statement

Data are restricted due to the limited permissions given by the participants.

## Ethical approval

Ethical approval was granted by the Faculty of Medicine and Health Sciences Research Committee at the University of Nottingham (reference FHMS 39-1023).

## Focus group schedule for children and young people on raising the age of sale of tobacco in the UK

The questions are mapped across to the COM-B framework: C – capability, O – opportunity, M – motivation.

### Before interview

Explain purpose of study. Go through the participant information sheet with participants. Reiterate points about consent, confidentiality, safeguarding, recording, treatment of data and withdrawal. Remind them that everything said is confidential and should not be repeated outside of the room.

**Interviewer: Do you have any questions about any part of the study?**

### Personal experiences of smoking/vaping

**1.** Does anybody know someone who smokes or vapes? **Probe: who, do they know how they started?**

**2.** Has anybody tried smoking or vaping themselves? **Probe: how did you start?**

**3.** What do you think are the main reasons that young people start smoking?

### Probe: peer pressure, (stress/boredom), control/rebellion?

**4.** How do you or people you currently know get hold of cigarettes or vapes? **(O, C)**

### Concept of smokefree generation law (SFG)

**5.** Do you know what the current age of sale of cigarettes is in the UK? **(C)**

**6.** Do you know what the current age of sale for vapes/e-cigarettes is in the UK? (C)

*Explain that it is currently 18. Explain that the Government has a plan to change this in England. Shops and other sellers could be banned from ever selling to anyone born after a certain year e.g. to children who are currently 14 (those born in 2009 or later). So, even when 14-year-olds reach 18, 19, 20, they wouldn’t be able to buy cigarettes. Individuals could not be criminalised or punished for buying cigarettes – only shops could be punished for selling them. This would raise the age of sale one year every year. The age of sale for vapes would remain at 18*.

**7.** Do you understand the law? **(C)**

**8.** What are your thoughts on this idea if it were introduced?

**9.** Think about people who currently smoke but want to quit. What would this measure have meant for them? **(O, C, M)**

**10.** What are your thoughts on whether this could stop young people from trying a cigarette in the first place? **(O, C) Probe: How might they get hold of a cigarette?**

**11.** What are your thoughts on whether this could stop young people from trying an e-cigarette in the first place? **(O, C, M)**

### Implementation of SFG

**12.** Do you have any other thoughts around how age of sale laws might work?

**13.** How might these kinds of new laws affect how young people think about cigarettes? **(M)**

**14.** How might these kinds of new laws affect how young people think about vapes? **(M)**

### Probe: Do you think it would change how young people talk about cigarettes/vapes with their friends? (M)

**15.** How might this law change how cigarettes/vapes are portrayed in the media or social media you use? **(M)**

### Probe: Are there any other negatives or benefits to the law?

**16.** What would it be like to be part of a generation where nearly no-one smokes? (M)

### What do you think of the following ideas?

**17.** The Government sets up a group of young people to advise them on how to implement the Smokefree Generation idea. **(O, C, M)**

### PROBE: What sort of young people should be on the group? How much influence should they have?

**18.** The Government introduces licenses for retailers who still sell tobacco to young people. If they are caught selling tobacco to those underage, they lose their license. **(O, C)**

### PROBE: How effective would that be?

**19.** The government runs a marketing campaign to inform people about the age of sale law. **(C, M)**

### PROBE: What would be the most important messages? Who should communicate these? Should there be different messages for different groups?

**20.** Are there other rules, laws or support, if any, do you think should be put in place at the same time as new laws on age of sale? **(O, C, M)**

**21.** Do you have any other thoughts you’d like to share?

Thank participants for their participation and explain the next steps for the research.

